# Lower prevalence of Post-Covid-19 Condition following Omicron SARS-CoV-2 infection

**DOI:** 10.1101/2023.04.05.23288157

**Authors:** Siméon de Bruijn, Albert Jan van Hoek, Elizabeth N. Mutubuki, Hans Knoop, Jaap Slootweg, Anna D. Tulen, Eelco Franz, Cees C. van den Wijngaard, Tessa van der Maaden

## Abstract

**Background:** Different SARS-CoV-2 variants can differentially affect the prevalence of Post Covid-19 Condition (PCC). This prospective study assesses prevalence and severity of symptoms three months after an Omicron infection, compared to Delta, test-negative and population controls. This study also assesses symptomology after reinfection and breakthrough infections.

**Methods:** After a positive SARS-CoV-2 test, cases were classified as Omicron or Delta based on ≥ 85% surveillance prevalence. Population controls were representatively invited and symptomatic test-negative controls enrolled after a negative SARS-CoV-2 test. Three months after enrolment, participants indicated point prevalence for 41 symptoms and severity of four symptoms. Permutation tests identified significantly elevated symptoms in cases compared to controls. PCC prevalence was estimated as the difference in prevalence of at least one elevated symptom in cases compared to population controls.

**Findings:** At three months follow-up, five symptoms and severe dyspnea were significantly elevated in Omicron cases (n = 4138) compared to test-negative (n= 1672) and population controls (n= 2762). PCC prevalence was 10·4% for Omicron cases and 17·7% for Delta cases (n = 6855). Prevalence of severe fatigue and dyspnea were higher in reinfected compared to primary infected Omicron cases, while severity of symptoms did not significantly differ between Omicron cases with a booster or primary vaccination course.

**Interpretation:** Three months after Omicron, prevalence of PCC is 41% lower than after Delta. Reinfection seems associated with more prevalent severe long-term symptoms compared to a first infection. A booster prior to infection does not seem to improve the outcome of long-term symptoms.

**Funding:** The study is executed by the National Institute for Public Health and the Environment by order of the Ministry of Health, Welfare and Sport.

## INTRODUCTION

Worldwide an estimated 750 million SARS-CoV-2 infections have occurred up to March 2023, and numerous publications report that in a subgroup of COVID-19 infected individuals symptoms persist for months (1-3). This condition is commonly referred to as Post Covid-19 Condition (PCC), which can burden health care systems and have a significant impact on individuals (3). A case definition of PCC by the WHO stipulated a general difficulty functioning in everyday life (4). However, prevalence and severity of symptoms associated with PCC may vary with different variants of SARS-CoV-2 (5). Compared to B.1.617.2 (Delta), B.1.1.529 (Omicron) has already been characterized by higher transmissibility, lower pathogenicity and a shorter duration of the acute phase (6). Moreover, acute symptoms differ with less involvement of the lower respiratory tract (6). Although it can be hypothesized that milder infections lower the risk of PCC, information on the prevalence and severity of symptoms associated with PCC after Omicron infections is still limited. Additionally, Omicron is better at immune escape than Delta which also raises the question to what extent vaccination protects against PCC-related symptoms after Omicron breakthrough infections and to what extent a previous infection may protect against PCC-related symptoms after an Omicron reinfection (7).

For pre-Omicron variants of concern, the prevalence and severity of these sequela are already documented for several countries including the Netherlands (2, 8-10). In the Netherlands, over 8·5 million SARS-CoV-2 infections were reported up to March 2023 since the onset of the pandemic, of those nearly 4·2 million infections occurred from December 2021 on during the Omicron variant of concern emergence (11).

This study aimed to assess PCC symptom prevalence and severity after infections with the Omicron variant compared to a Delta infection, test-negative controls and population controls. Moreover we assessed the possible protective effect of the booster vaccination against developing PCC-related symptoms after Omicron breakthrough infections. Finally we investigated the prevalence of PCC-related symptoms after a first infection with Omicron compared to those with a reinfection with Omicron after a preceding COVID-19 infection.

## METHODS

### Design, participants and inclusion

Data were collected in the context of the national Dutch prospective LongCOVID-study. Study design details are described in the previously published study protocol (12). This paper reports a follow-up study from our previous findings on long term prevalence and severity of symptoms 3 months after Alpha and Delta SARS-CoV-2 infection (8). In brief, here we report on Omicron cases aged 18 or older three months after testing positive for SARS-CoV-2 and were enrolled between January 3^rd^ and May 31^st^ 2022 **(Figure 1)**. The start of the Omicron dominant period is defined by ≥ 85% prevalence of Omicron in the Dutch pathogen surveillance (13). Likewise we included Delta cases that enrolled during ≥ 85% prevalence pathogen prevalence between July 5^th^ 2021 and December 19^th^ 2021. Cases between December 19^th^ 2021 and January 3^rd^ were excluded as neither Delta nor Omicron was dominant during this period in the Netherlands. Cases were recruited, as defined in more detail in the study protocol, within seven days following a positive polymerase chain reaction or self- or professionally administered rapid lateral flow SARS-CoV-2 test. Test-negative controls who reported respiratory symptoms as reason to test for SARS-CoV-2 and population controls without previous suspected or confirmed COVID-19 were included to control for background prevalence of long term symptoms. Population controls from the Netherlands were randomly invited by direct mailing. Both control groups were included if they enrolled between July 5^th^ 2021 and May 31^st^ 2022. Participants could voluntarily self-register on the study’s website (longcovid.rivm.nl) and received questionnaires at baseline (T0) and after three months follow-up (T3).

**Figure 1:**
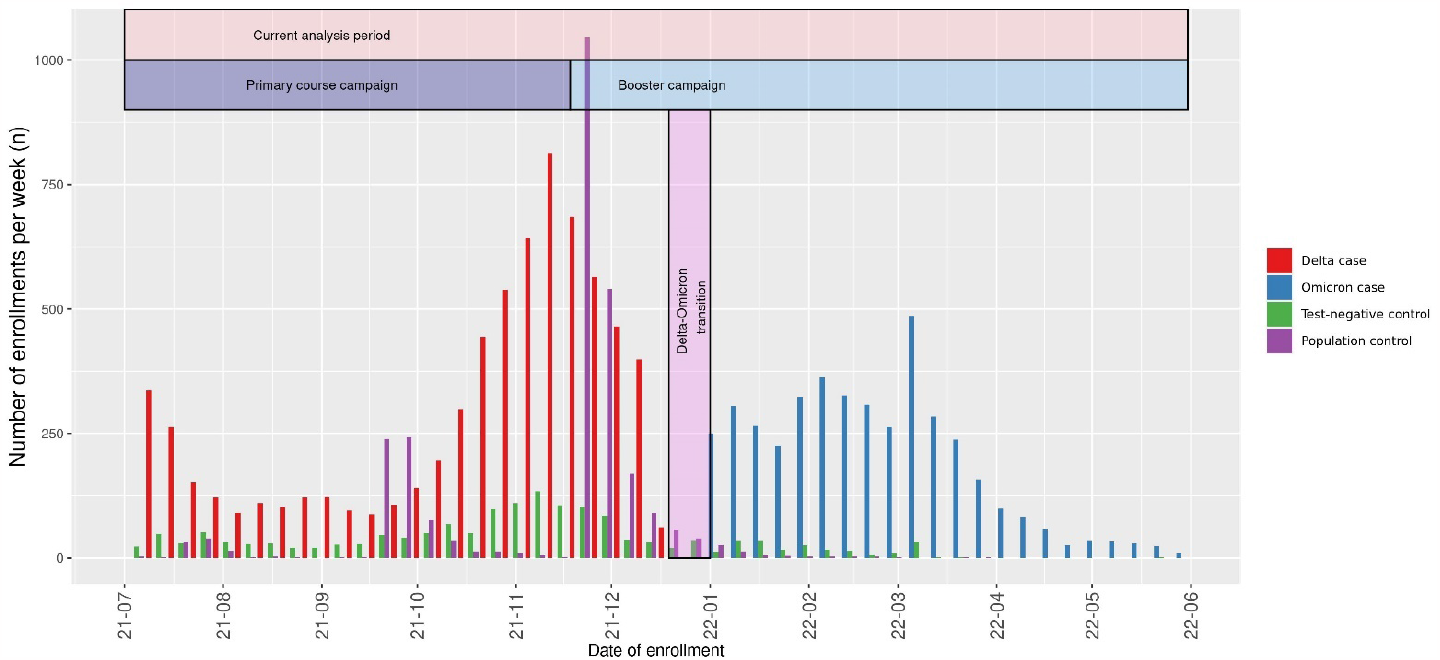
Timeline of, the current analysis period, the Dutch COVID-19 vaccination program and inclusion and classification of participants in the current study.

### Outcomes and covariates

The primary outcome was the prevalence of PCC, which we defined – slightly adapted from our previous study – as the difference in prevalence of at least one significantly elevated symptom in Omicron cases compared to the prevalence in the population control group after three months (8). Likewise we assessed the prevalence of PCC in Delta cases. As secondary outcomes we assessed the prevalence of symptoms with a clinically relevant severity, using validated questionnaires with population norm scores. These included severe fatigue measured with the subscale fatigue of the Checklist Individual Strength (CIS, cut-off score ≥ 35) (14, 15); severe self-reported cognitive problems on the Cognitive Failure Questionnaire (CFQ, cut-off score ≥ 44) (16, 17); severe pain on the bodily subscale of the RAND SF-36 Health Status Inventory (SF-36, cut-off score ≤55) (18); and severe dyspnea on the modified Medical Research Council (mMRC, cut-off score ≥1) scale (19). We compared the prevalence of severe fatigue, severe cognitive problems, severe pain and severe dyspnea for Omicron cases, Delta cases, test-negative controls and population controls. Furthermore we analysed the prevalence of severe symptoms in Omicron cases between a group that received a booster vaccination and a group that only completed a primary course. A completed primary course was defined as having received two doses of the COVID-19 vaccine at least 14 days prior to a positive SARS-CoV-2 test, or one dose of JCOVDEN at least 28 days prior, or reporting a preceding suspected or confirmed COVID-19 infection and subsequently receiving one dose at least 14 days prior to a baseline positive SARS-CoV-2 test. A booster was defined as having received an additional dose after a complete primary course (for details see **Supplementary methods**). Finally, we analysed the prevalence of severe symptoms in Omicron cases between a first infection group without a previous SARS-CoV-2 infection and a reinfection group with a previous suspected or confirmed infection. In the reinfection group, the prior infection could be either suspected or test-confirmed, since in the early phase of the pandemic testing was not sufficiently available yet. Reinfected cases were excluded if at T0 they self-reported long-term symptoms which they attributed to their prior infection.

Information on demographics, vaccination status, general health status, use of health care and medication, and comorbidities (adapted from the TiC-P) (20) were collected at baseline.

### Statistical analyses

Statistical procedures were based on a predefined, published study protocol (12). Briefly, the primary analysis was a complete case analysis with only participants completing both T0 and T3. Four additional sensitivity analyses were used to substitute for missing data on symptoms at T3: multiple imputation, carry forward, best case and worst case scenario (See **Supplementary Methods**).

Prevalence of 41 symptoms and prevalence of severe fatigue, severe cognitive impairment, severe pain and severe dyspnea were compared between Omicron cases, Delta cases and both control groups by permutation tests which stratified for predefined confounders age, sex, level of education and number of comorbidities. Significantly elevated symptoms in Omicron cases compared to controls were defined by a two-sided 5% significance level with p-values adjusted for multiple testing by the Benjamini-Hochberg procedure (21). Prevalence of at least one significantly elevated symptom at T3 was then assessed for Omicron and Delta cases and both control groups. Prevalence of PCC was then estimated by the difference in prevalence of at least one significantly elevated symptom in the cases compared to the population controls. Likewise, comparisons were made for Omicron cases with a booster and with only a completed primary vaccination course and for Omicron cases with a first infection and a reinfection. Lastly, to evaluate the sensitivity of the PCC definition we compared the severity scores between cases and population controls who did not have one of the significantly elevated symptoms at T3.

Analyses were performed with R version 4·1·0 (packages listed in **Supplementary methods**).

### Ethics approval

In February 2021 the Utrecht Medical Ethics Committee (METC) declared that the Medical Research Involving Human Subjects Act (WMO) does not apply to this study as it is survey based (protocol number 21-124/C).

## RESULTS

Baseline characteristics for cases and both control groups that completed both T0 and T3 questionnaires (complete case) are shown in **Table 1**. In total 4138 Omicron cases, 6855 Delta cases, 1672 test-negative controls and 2726 population controls are included. Differences between controls and cases in vaccination status are mostly due to differing inclusion times. Additionally, population control generally have less comorbidities and a lower education compared to cases.

**Table 1.** Demographics and acute illness at baseline

| Complete case | Omicron cases | Delta cases | Test-negative controls | Population controls |
| --- | --- | --- | --- | --- |
| N | 4138 | 6855 | 1672 | 2726 |
| Age, median [IQR] | 55.8 [43.8 ; 65.9] | 52.1 [40.0 ; 62.8] | 57.3 [44.0 ; 66.0] | 53.2 [41.9 ; 60.7] |
| Sex, % (n) |  |  |  |  |
| Female | 62.0 (2567) | 62.9 (4315) | 64.4 (1076) | 68.6 (1871) |
| Male | 37.7 (1561) | 36.8 (2526) | 35.1 (587) | 31.2 (850) |
| Other | 0.1 (4) | 0.1 (9) | 0.4 (6) | 0.0 (1) |
| Pregnancy, % (n) | 2.2 (19) | 2.4 (45) | 2.1 (7) | 4.1 (27) |
| BMI, mean (SD) | 25.91 (4.68) | 25.76 (4.60) | 26.00 (4.86) | 25.84 (4.65) |
| Smoking, % (n) |  |  |  |  |
| Current smoker | 3.9 (160) | 4.4 (301) | 7.4 (123) | 5.7 (156) |
| Former smoker | 27.0 (1117) | 25.6 (1754) | 30.1 (503) | 21.2 (577) |
| Never smoker | 67.0 (2771) | 67.3 (4614) | 59.7 (999) | 71.0 (1935) |
| Level of education, % (n) |  |  |  |  |
| Low | 3.2 (132) | 3.4 (235) | 2.2 (37) | 5.2 (142) |
| Medium | 32.0 (1324) | 35.0 (2402) | 26.6 (445) | 38.5 (1049) |
| High | 64.8 (2682) | 61.5 (4218) | 71.2 (1190) | 56.3 (1535) |
| History with COVID-19, % (n) | 10.9 (453) | 9.0 (617) | 0.0 (0) | 0.0 (0) |
| Nr of comorbidities, % (n) |  |  |  |  |
| 0 | 44.2 (1827) | 47.1 (3231) | 40.0 (668) | 54.0 (1471) |
| 1-2 | 42.7 (1766) | 42.3 (2902) | 42.9 (718) | 36.5 (996) |
| >2 | 13.2 (545) | 10.5 (722) | 17.1 (286) | 9.5 (259) |
| Respiratory disease, % (n) | 17.4 (720) | 16.6 (1137) | 21.5 (359) | 11.3 (309) |
| Hypertension, % (n) | 14.1 (584) | 12.0 (826) | 14.8 (248) | 10.8 (294) |
| Diabetes, % (n) | 3.7 (155) | 2.9 (202) | 3.8 (63) | 3.4 (93) |
| Cardiovascular disease, % (n) | 2.5 (102) | 2.0 (135) | 2.8 (47) | 1.4 (38) |
| Use of healthcare, % (n) | 6.2 (257) | 10.6 (727) | 12.0 (200) | 5.5 (151) |
| Medication use, % (n) | 74.5 (3083) | 77.2 (5292) | 68.7 (1148) | 22.2 (605) |
| Admitted to hospital, % (n) | 0.2 (7) | 0.1 (5) | 0.1 (1) | 0.5 (6) |
| Vaccination status at T0, % (n) |  |  |  |  |
| Boostered | 76.1 (2970) | 0.3 (18) | 11.0 (160) | 1.9 (46) |
| Complete primary course | 21.8 (853) | 93.4 (5274) | 83.1 (1206) | 94.9 (2346) |
| Partially vaccinated | 0.5 (21) | 2.1 (118) | 3.3 (48) | 1.5 (37) |
| Unvaccinated | 1.5 (60) | 4.2 (238) | 2.5 (37) | 1.7 (43) |
| Number of symptoms at T0, median [IQR] | 8 [5; 12] | 9 [6; 13] | 5 [3;8] | 0 [0; 2] |
| Variant dominant at T0, % (n) |  |  |  |  |
| Delta | 0.0 (0) | 100 (6855) | 83.0 (1250) | 94.6 (2551) |
| Delta-Omicron | 0.0 (0) | 0.0 (0) | 3.8 (57) | 3.2 (87) |
| Omicron | 100 (4138) | 0.0 (0) | 13.2 (199) | 2.2 (58) |

**Figure 2** shows that fatigue (24·3%; p.BH = 0·0077), dyspnea (11·1%; p.BH = 0·025) difficulties with a busy environment (8·4%; p.BH = 0·0077), problems with memory (5·9%; p.BH = 0·00092) and brainfog (2·9%; p.BH = 0·014) were significantly elevated in Omicron cases compared to both control groups (shown p.BH-values here are compared to test-negative controls, all p.BH < 0·0001 compared to population controls) after three months follow-up in the complete case scenario. Yet, the prevalence of all five symptoms was significantly lower in Omicron cases compared to Delta cases. Prevalence of all 41 symptoms per group for T0 and T3 are available in **Supplementary Table S1**.

**Figure 2:**
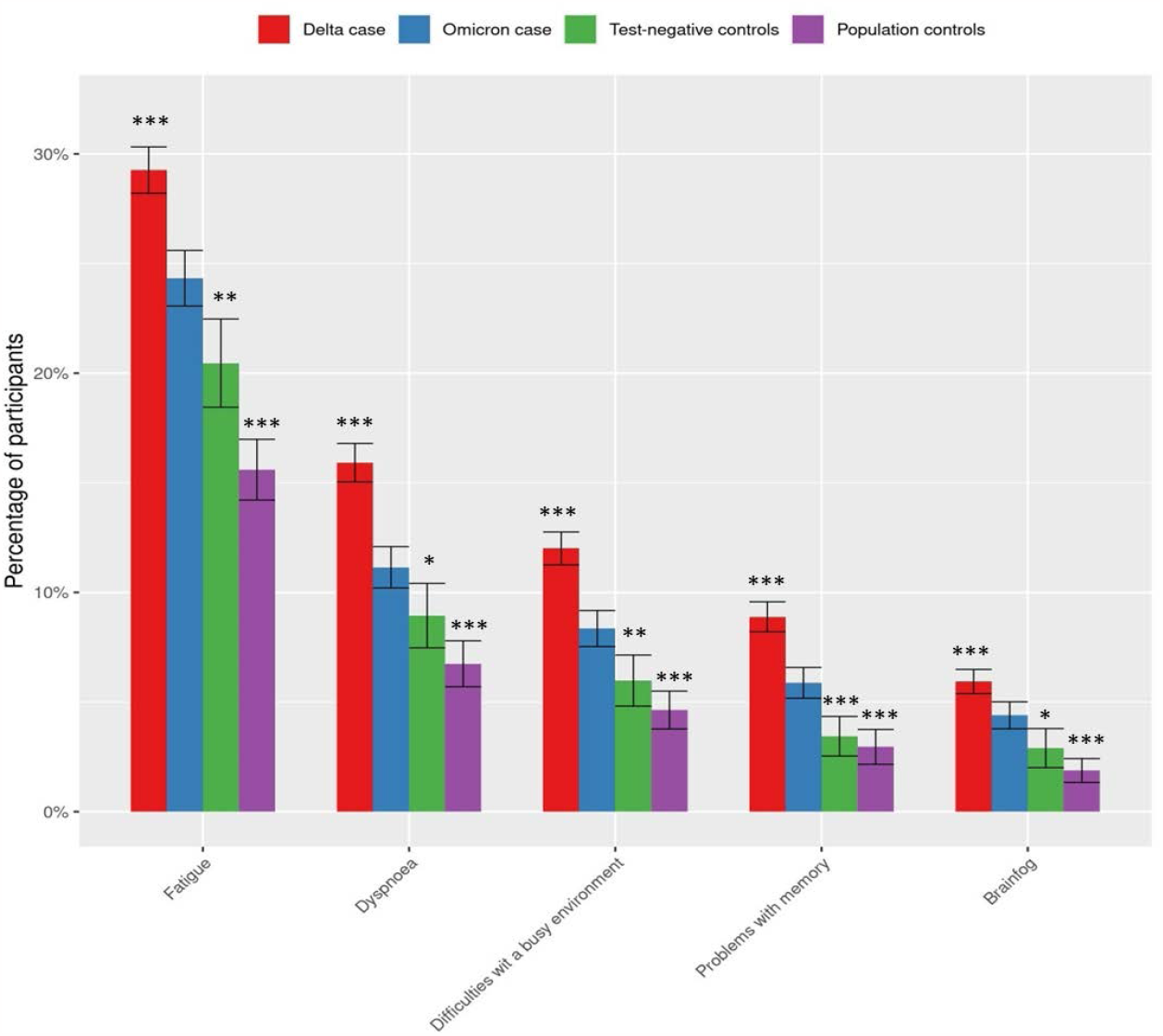
Standardised prevalence (95% confidence intervals) of the 5 symptoms at T3 that were significantly elevated (p.BH<0,05) between Omicron cases and both control groups and their prevalence in Delta cases using complete case analysis without substituting for missing values at T3. Symptoms are ranked by prevalence in Omicron cases. *BH.adjusted p-value <0.05; **BH.adjusted p-value <0.01 ; ***BH.adjusted p-value <0.001 compared to Omicron cases.

Of the significantly elevated symptoms, fatigue and dyspnea were generally reported in both the acute phase and at T3 while difficulty with a busy environment, problems with memory and brainfog were generally reported more at T3 only **(Figure S2)**.

Severe dyspnea (mMRC score≥1) had a significantly higher prevalence in Omicron cases (8·3% prevalence) compared to test-negative and population control groups (6·3% and 4·2% respectively, with p.BH 0·02 and < 0·001); **Figure 3**). Severe fatigue (CIS score ≥ 35) and severe cognitive problems (CFQ score ≥ 44) were significantly more prevalent in Omicron cases (11·8%) compared to population controls (6,6%, p.BH < 0·001) but not compared to test-negative controls (10·6%, p.BH = 0·14). Finally, severe fatigue (25·6% vs 22·1%), severe cognitive problems (14·3% vs 11·8%) and severe dyspnea (12·2% vs 8·3%) were significantly more prevalent in Delta cases compared to Omicron cases (all p.BH <0·0001).

**Figure 3:**
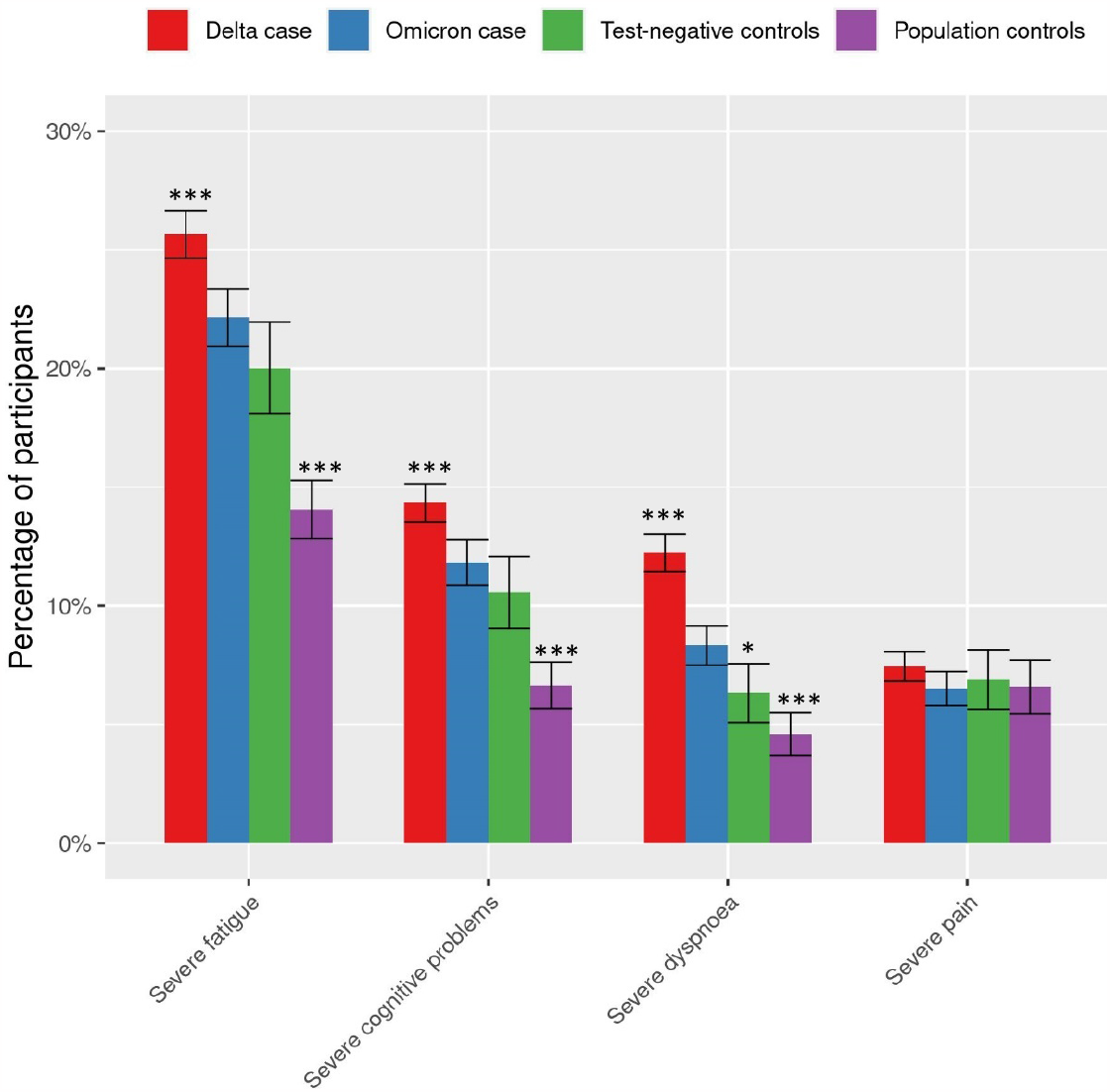
Standardised prevalence (95% confidence intervals) of severity score cut-off values in cases and both control groups using complete case analysis without substituting for missing values at T3. Severe fatigue: Checklist Individual Strength (CIS), subscale fatigue ≥35, severe cognitive problems: Cognitive Failure Questionnaire (CFQ) ≥44, severe dyspnoea: modified Medical Research Council dyspnoea scale mMRC ≥1, severe pain: SF-36 subscale bodily pain ≤55. *BH.adjusted p-value <0.05; **BH.adjusted p-value <0.01 ; ***BH.adjusted p-value <0.001 compared to Omicron cases.

In the complete case scenario, prevalence of PCC, i.e. the difference in prevalence of at least one of the five significantly elevated symptoms in cases compared to the population controls, was estimated at 10·4% in Omicron cases, compared to 17·7% in Delta cases. The overall prevalence of the five significantly elevated symptoms was found to be significantly lower for Omicron cases (30·0%) compared to Delta cases (37·2%) but higher compared to test-negative controls (26·2%) and population controls (19·6%) at T3 **(Figure 4)**. Differences between Omicron cases and Delta cases and control groups were also noticeable in the multiple imputation, carry forward and worst case scenario when substituting for missing values **(Supplementary Figure S1)**. In the carry forward scenario prevalence of PCC was higher largely due to differences in cases and population controls at T0, where cases and test-negative controls were in the acute phase of an infection. In the best case scenario differences between Omicron cases and controls were no longer significant, likely due to the substitution of missing values at T3 with complete recovery, but differences between Omicron and Delta were still present.

**Figure 4:**
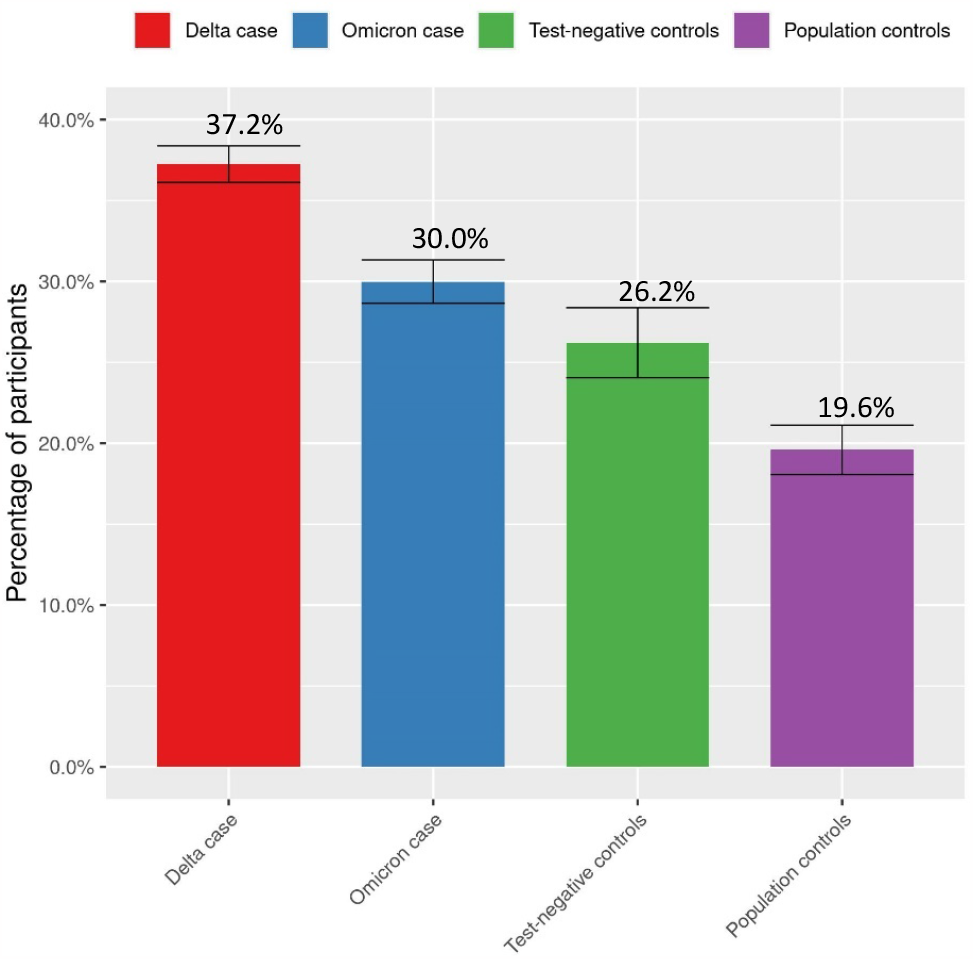
Standardized prevalence (95% confidence intervals) of at least one of the five significantly elevated symptoms at T3 in Delta and Omicron cases compared to test-negative and population controls.

Of the Omicron cases, in the complete case analysis 10·9% (453 out of 4138) of the participants reported a suspected or confirmed prior SARS-CoV-2 infection before enrolment. Of the reinfected cases, 438 had one infection prior to their inclusion Omicron infection and 15 had two or more. Baseline demographics are available in Supplementary Table S3. Severe fatigue (28·0% ; p.BH = 0·030) and severe dyspnea (12·1%; p.BH = 0·020) were significantly more prevalent in Omicron cases with a reinfection compared to Omicron cases with a first SARS-CoV-2 infection (23·0% and 8·1% respectively). Additionally, of the five significantly elevated symptoms, here too fatigue (p.BH = 0·014) and dyspnea (p.BH = 0·045) were reported more frequently in the reinfection group **(Supplementary Figure S3)**. Prevalence of at least one significantly elevated symptom was 37·8% for reinfected cases and 30·3% for cases with a first infection.

Of the Omicron cases, 2970 (76·1%) participants received a booster vaccination while 853 (21·8%) participants only completed the primary course prior to infection in the complete case analysis. Baseline demographics are available in **Supplementary Table S2**. Severe fatigue and severe cognitive problems were less prevalent in cases with a booster (21·0% and 10·9% respectively) compared to the primary course cases (23·1% and 13·2%), though neither significantly **(Supplementary Figure S3)**.

Prevalence of at least one of the five significantly elevated symptoms was similar for boostered cases and cases with a completed primary course (27·4% and 28·8% respectively). None of the five PCC-related symptoms were significantly different in prevalence between the two groups **(Supplementary Figure S3)**. There were too few partially vaccinated cases (n = 21; 0·5%) and unvaccinated cases (n = 60; 1·5%) to analyse. Cases that did not fulfil our case definition had significantly worse scores for CIS-fatigue, CFQ and SF36-pain than population controls not fulfilling the case definition, but the absolute differences were less than 1 point on all three scales **(Supplementary Table S4)**.

## DISCUSSION

In this prospective cohort study we found that three months after an Omicron infection, symptoms that were significantly elevated compared to both control groups were fatigue, dyspnea, difficulties with a busy environment, problems with memory and brainfog. Prevalence of PCC – i.e. the difference in prevalence of at least one of these symptoms in cases compared to the population controls – was 41% lower for Omicron cases compared to Delta cases: prevalence of at least one symptom possibly associated with PCC (30·0%) was lower compared to a Delta infection (37·2%) yet still exceeding the background prevalence in population controls (19·6%).

Severity of symptoms was also lower after Omicron than after Delta for fatigue, cognitive impairment and dyspnea. Still these symptoms were reported significantly more often as severe in Omicron cases compared to population controls, whereas only severe dyspnea was increased compared to test-negative controls. Previous research in the same study on three month follow-up of prevalence and severity of symptoms following an Alpha or Delta infection indicated a total of 13 significantly elevated symptoms (8). In the Omicron analysis only 5 symptoms exceeded background prevalence in both control groups. Most notably, the COVID-19 characteristic symptoms of loss of smell and loss of taste are 5·5 and 4·2 times higher in prevalence at T3 for Delta cases than Omicron cases **(Supplementary Table 1)**. A possible explanation for these findings could be that Omicron generally has less involvement of the lower respiratory tract and a milder acute phase than Delta (6). These findings are in line with a Danish and UK study that have shown lower odds for PCC with Omicron compared to Delta (22, 23). Studies have shown that PCC is associated with the severity of the acute phase of COVID-19 (24, 25). In the current study we only find that the median number of symptoms at T0 is slightly higher for Delta cases compared Omicron cases, but considerably more compared to both control groups **(Table 1)**. However, prevalence of severe fatigue and severe bodily pain was significantly higher in the acute phase for Delta cases than Omicron cases. Interestingly, a Norwegian study did not find differences in long-term symptoms after Omicron and Delta infections when the variants co-circulated (26). This discrepancy with our and other studies may have to do with different levels of immunity in the population when comparing Omicron and Delta infections in different calendar study periods or differences in subvariants analysed.

Cases with a reinfection seemed to have a higher prevalence for PCC with a 1·2 times higher prevalence of PCC-related symptoms compared to a first SARS-CoV-2 infection with the Omicron variant. In fact, the prevalence of at least one of the significantly elevated symptoms at T3 amongst reinfected cases was very similar to the prevalence in the Delta case group (37·8% and 37·2% respectively). Estimates suggest over half a billion people globally have been infected SARS-CoV-2, which infers an increasing likelihood of reinfection occurring with (sub)variants more adept at immune escape. Research has shown that Omicron is indeed associated with a marked ability to evade immunity from prior infection (27). Natural immunity, and also hybrid immunity with vaccination, against subsequent infection has additionally been shown to wane over time (28). However, it cannot be excluded that reinfected cases had impaired health possibly putting them at higher risk both for reinfection and PCC. This would imply that the probability of developing symptomatic COVID-19 due to different health status rather than the reinfection itself would somehow be associated with a higher risk of developing PCC. This would be in line with another study that has shown that hybrid immunity from prior infection and vaccination did not abrogate risk of long-term symptoms (29). Additional research on PCC and reinfection is needed to better understand the dynamics that govern these findings.

Waning immunity may also infer a diminishing effect of vaccination on protection against PCC after an Omicron infection. Omicron cases with a primary course received their last vaccination a median 142 days prior to infection while boostered cases had a median time difference of 60 days. Still the booster vaccinations compared to a primary vaccination course seemed at most modestly protective for PCC: fatigue and cognitive problems were less frequently severe in cases with a booster, but these differences were not statistically significant and the study may lack the power to detect prevalence differences smaller than 5% (12). Likewise, prevalence of at least one possible PCC-related symptom was similar between both groups. Studies have shown a partial protection of the primary course compared to unvaccinated cases for pre-Omicron variants (8, 30, 31) and for Omicron (32). Research on the effect of the booster is limited but one study shows a lower association with PCC for three-dose vaccinated Omicron cases compared to two-dosed (23). Generally, there is some evidence to suggests that a booster vaccination provides an albeit temporary protection against infection with Omicron (33). Indirect effects by preventing infection and transmission compounded with a modest direct effect may still yield a more than modest reduction in PCC incidence. Currently, bivalent COVID-19 vaccines are available which contain, in addition to the wild-type SARS-CoV-2 mRNA, Omicron BA·1 or Omicron BA·4-5 subvariants mRNA. Possibly, these bivalent vaccines may infer increased protection against PCC. Still, our findings suggest that monovalent booster vaccine induced immunity has either waned or offers limited direct protection against long-term symptoms following an Omicron breakthrough infection.

### Strengths and limitations

A strength of this prospective cohort study is the inclusion of large numbers of Delta and Omicron cases as well as two control groups to be able to estimate the prevalence of PCC corrected for the background prevalence of symptoms in the population and symptoms likely due to other respiratory infections. Reporting of reinfections and changing vaccination status during follow up made it possible to investigate their association with the prevalence of PCC after SARS-CoV-2 infection. Moreover, recruiting through test sites rather than through hospitals likely resulted in a cohort that is representative of the long-term effects of COVID-19 in the general population.

This study also has some limitations. Firstly, data was collected exclusively through online questionnaires without clinical evaluation of symptoms. Nevertheless our use of validated questionnaires with population norm scores and the aforementioned control groups made it possible to assess to what extent prevalence of reported symptoms exceeded the background prevalence. Secondly, the follow-up of the T3 survey had a response rate of 70%. It is possible that the missing 30% dropped-out due to lack of symptoms or, oppositely, becoming severely ill. Therefore we substituted for missing values by multiple alternative imputation scenarios, which showed robustness of our finding that Omicron was less severe than Delta. Thirdly, recruitment of controls during the Omicron period was low. Background prevalence was therefore largely established on controls recruited during the Delta period which may have a different background prevalence than the Omicron period due to different COVID-19 restrictions and seasonal effects. However, most controls included during the Delta period were enrolled closely to the start of the Omicron period with only a 40 day median difference. **Supplementary Table 1** additionally shows that between T0 and T3 background prevalence of symptoms in population controls fluctuates at most with 3,1 percentage point, indicating little fluctuation over time. Moreover, severe fatigue from our control groups (20·0% for test-negative and 14·0% for the population control) was similar to a large Dutch population cohort (18%, n=78363)(34). As Delta circulated during summer and autumn in the Netherlands and Omicron has, at the moment of analysis, circulated during winter and spring, correction for seasonal effects between Delta and Omicron cases was not possible. This means that some symptoms that significantly exceeded the background prevalence for Delta cases but not for Omicron cases might be explained by other factors than the variant. Variant attribution was performed on 85% Dutch pathogen surveillance proportion, which may lead to marginal misclassification for the weeks of transition between Alpha and Delta and between Delta and Omicron. As well, subvariants BA·1 and BA·2 of Omicron have circulated in the Netherlands during the study period (13). Due to the relatively long co-circulation of subvariants, and the absence of SARS-CoV-2 PCR test sequence data, subvariant specific analysis are not possible. Finally, our case definition of PCC may not have been sensitive enough to capture all post-covid symptoms, since cases that did not fulfil our case definition had significantly worse scores for CIS-fatigue, CFQ and SF36-pain than population controls not fulfilling the case definition. However, the absolute differences in mean scores were at most minimal, see **Supplementary Table S4)**, which suggests that our case definition still captured the vast majority of PCC.

### Implications

These findings suggest that the prevalence of PCC is two fifths lower following an Omicron infection compared to Delta. Reinfection appears to be associated with more prevalent long-term symptoms compared to a first infection. A preceding booster vaccination does not seem to improve the outcome regarding PCC in Omicron cases.

## Supporting information

Supplementary Methods and Results

## Data Availability

Supporting clinical documents including the study protocol and statistical analysis plan will be available immediately following publication of this Article for at least 1 year. Researchers who provide a methodologically sound proposal will within the applicable privacy legislation be allowed to access to the de-identified individual participant data that underlie the results reported in this article. Proposals should be sent to the corresponding author. These proposals will be reviewed and approved by the investigators on the basis of scientific merit. To gain access, data requestors will need to sign a data access agreement.

## DECLARATIONS

### List of abbreviations

CIS: Checklist Individual Strength
CFQ: Cognitive Failure Questionnaire
mMRC: Modified Medical Research Council dyspnea scale
PCC: Post-Covid-19 Condition
PCR: polymerase chain reaction
SARS-CoV-2: Severe acute respiratory syndrome coronavirus 2
SF-36: SF-36 item Health Survey
TiC-P: Treatment Inventory of Costs in Patients with psychiatric disorders

## Author’s contributions

TM, AH, CW and EF conceptualised the study. EM, CW, HK designed the study protocol and statistical analysis. SB, TM and JS analysed the data. SB, TM, EM, AT, AH, EF and CW contributed to the data interpretation. SDB and TM coordinated the data collection and SDB drafted the manuscript. All authors reviewed and edited revisions of the manuscript, had full access to all the data in the study, and had final responsibility for the decision to submit for publication

## Acknowledgements

We thank Caroline van den Ende for structural updating of literature on PCC.

## REFERENCES

1. World Health Organization. 2021. WHO coronavirus (COVID-19) dashboard. Access date 4 May 2022. [Available from: https://covid19.who.int/.

2. Groff D, Sun A, Ssentongo AE, Ba DM, Parsons N, Poudel GR, et al. Short-term and Long-term Rates of Postacute Sequelae of SARS-CoV-2 Infection: A Systematic Review. JAMA Netw Open. 2021;4(10):e2128568.

3. Parums DV. Editorial: Long COVID, or Post-COVID Syndrome, and the Global Impact on Health Care. Med Sci Monit. 2021;27.

4. Soriano JB, Murthy S, Marshall JC, Relan P, Diaz JV. A clinical case definition of post-COVID-19 condition by a Delphi consensus. Lancet Infect Dis. 2021.

5. Fernández-de-las-Peñas C, Notarte KI, Peligro PJ, Velasco JV, Ocampo MJ, Henry BM, et al. Long-COVID Symptoms in Individuals Infected with Different SARS-CoV-2 Variants of Concern: A Systematic Review of the Literature. Viruses. 2022;14(12):2629.

6. Menni C, Valdes AM, Polidori L, Antonelli M, Penamakuri S, Nogal A, et al. Symptom prevalence, duration, and risk of hospital admission in individuals infected with SARS-CoV-2 during periods of omicron and delta variant dominance: a prospective observational study from the ZOE COVID Study. The Lancet. 2022;399(10335):1618–24.

7. Bowe B, Xie Y, Al-Aly Z. Acute and postacute sequelae associated with SARS-CoV-2 reinfection. Nat Med. 2022;28(11):2398–405.

8. van der Maaden T, Mutubuki EN, de Bruijn S, Leung KY, Knoop H, Slootweg J, et al. Prevalence and Severity of Symptoms 3 Months After Infection With SARS-CoV-2 Compared to Test-Negative and Population Controls in the Netherlands. The Journal of Infectious Diseases. 2022:1537–6613 (Electronic).

9. Ballering AV, van Zon SKR, Olde Hartman TC, Rosmalen JGM. Persistence of somatic symptoms after COVID-19 in the Netherlands: an observational cohort study. (1474-547X (Electronic)).

10. Chen C, Haupert SR, Zimmermann L, Shi X, Fritsche LG, Mukherjee B. Global Prevalence of Post COVID-19 Condition or Long COVID: A Meta-Analysis and Systematic Review. J Infect Dis. 2022;226(9):1593–607.

11. Rijksinstituut voor Volksgezondheid en Milieu (RIVM). Covid-19 aantallen per gemeente per publicatiedatum. 2022 [Available from: https://data.rivm.nl/covid-19/.

12. Mutubuki EN, van der Maaden T, Leung KY, Wong A, Tulen AD, de Bruijn S, et al. Prevalence and determinants of persistent symptoms after infection with SARS-CoV-2: protocol for an observational cohort study (LongCOVID-study). BMJ Open. 2022;12(7):e062439.

13. Rijksinstituut voor Volksgezondheid en Milieu (RIVM). Covid-19 rapportage van SARS-CoV-2 varianten in Nederland via de aselecte steekproef van RT-PCR positieve monsters in de nationale kiemsurveillance. 2022 [Available from: https://data.rivm.nl/covid-19/.

14. Worm-Smeitink M, Gielissen M, Bloot L, van Laarhoven HWM, van Engelen BGM, van Riel P, et al. The assessment of fatigue: Psychometric qualities and norms for the Checklist individual strength. J Psychosom Res. 2017;98:40–6.

15. Vercoulen JH, Swanink CM, Fennis JF, Galama JM, van der Meer JW, Bleijenberg G. Dimensional assessment of chronic fatigue syndrome. J Psychosom Res. 1994;38(5):383–92.

16. Ponds R, Van Boxtel M, Jolles J. De cognitive failure questionnaire als maat voor subjectief cognitief functioneren. Tijdschrift voor Neuropsychologie. 2006(2):37–45.

17. Broadbent DE, Cooper PF, FitzGerald P, Parkes KR. The Cognitive Failures Questionnaire (CFQ) and its correlates. Br J Clin Psychol. 1982;21(1):1–16.

18. van der Zee KI, Sanderman R. Het meten van de algemene gezondheidstoestand met de RAND-36, een handleiding: umcg /Rijksuniversiteit groningen. Research Institute SHARE. 2012.

19. Mahler DA, Wells CK. Evaluation of clinical methods for rating dyspnea. Chest. 1988;93(3):580–6.

20. Bouwmans C, De Jong K, Timman R, Zijlstra-Vlasveld M, Van der Feltz-Cornelis C, Tan Swan S, et al. Feasibility, reliability and validity of a questionnaire on healthcare consumption and productivity loss in patients with a psychiatric disorder (TiC-P). BMC Health Serv Res. 2013;13:217.

21. Hochberg Y, Benjamini Y. More powerful procedures for multiple significance testing. Stat Med. 1990;9(7):811–8.

22. Antonelli M, Pujol JC, Spector TD, Ourselin S, Steves CJ. Risk of long COVID associated with delta versus omicron variants of SARS-CoV-2. Lancet. 2022;399(10343):2263–4.

23. Spiliopoulos L, Sørensen AIV, Bager P, Nielsen NM, Hansen JV, Koch A, et al. Post-acute symptoms four months after SARS-CoV-2 infection during the Omicron period: a nationwide Danish questionnaire study. medRxiv. 2022:2022·10·12·22280990.

24. Dryden M, Mudara C, Vika C, Blumberg L, Mayet N, Cohen C, et al. Post-COVID-19 condition 3 months after hospitalisation with SARS-CoV-2 in South Africa: a prospective cohort study. The Lancet Global Health. 2022;10(9):e1247–e56.

25. Iqbal FM, Lam K, Sounderajah V, Clarke JM, Ashrafian H, Darzi A. Characteristics and predictors of acute and chronic post-COVID syndrome: A systematic review and meta-analysis. EClinicalMedicine. 2021;36.

26. Magnusson K, Kristoffersen DT, Dell’Isola A, Kiadaliri A, Turkiewicz A, Runhaar J, et al. Post-covid medical complaints after SARS-CoV-2 Omicron vs Delta variants - a prospective cohort study. medRxiv. 2022:2022·05·23·22275445.

27. Pulliam JA-O, van Schalkwyk CA-O, Govender NA-O, von Gottberg AA-O, Cohen CA-O, Groome MA-O, et al. Increased risk of SARS-CoV-2 reinfection associated with emergence of Omicron in South Africa. (1095-9203 (Electronic)).

28. Goldberg Y, Mandel M, Bar-On YM, Bodenheimer O, Freedman LS, Ash N, et al. Protection and Waning of Natural and Hybrid Immunity to SARS-CoV-2. (1533-4406 (Electronic)).

29. Al-Aly Z, Bowe B, Xie Y. Outcomes of SARS-CoV-2 Reinfection. 2022.

30. Gao P, Liu JA-O, Liu MA-O. Effect of COVID-19 Vaccines on Reducing the Risk of Long COVID in the Real World: A Systematic Review and Meta-Analysis. LID - 10.3390/ijerph191912422 [doi] LID - 12422. (1660-4601 (Electronic)).

31. Al-Aly Z, Bowe B, Xie Y. Long COVID after breakthrough SARS-CoV-2 infection. Nat Med. 2022.

32. Ballouz T, Menges D, Kaufmann M, Amati R, Frei A, von Wyl V, et al. Post COVID-19 condition after Wildtype, Delta, and Omicron variant SARS-CoV-2 infection and vaccination: pooled analysis of two population-based cohorts. medRxiv. 2022:2022·09·25·22280333.

33. Ioannou GN, Bohnert ASB, O’Hare AM, Boyko EJ, Maciejewski ML, Smith VA, et al. Effectiveness of mRNA COVID-19 Vaccine Boosters Against Infection, Hospitalization, and Death: A Target Trial Emulation in the Omicron (B·1·1·529) Variant Era. Ann Intern Med. 2022.

34. Goërtz YMJ, Braamse AMJ, Spruit MA, Janssen DJA, Ebadi Z, Van Herck M, et al. Fatigue in patients with chronic disease: results from the population-based Lifelines Cohort Study. Sci Rep. 2021;11(1):20977.

