## Supplementary Methods and Results for "Lower prevalence of Post-Covid-19 Condition following Omicron SARS-CoV-2 infection"

### Supplementary files

**Table of contents:**

- Supplementary methods p2
- Supplementary results P4
- Table S1: Demographics and acute illness at baseline for complete cases (T0 and T3) P4
- Figure S1 p5
- Figure S2 p5
- Table S2. Demographics and acute illness at baseline for booster versus primary course cases P6
- Figure S3 p6
- Figure S4 p7
- Table S2. Demographics and acute illness at baseline for reinfected versus first infection cases P8
- Figure S5 p9
- Figure S6 p9
- Table S4. Severity score between non-PCC Omicron cases and non-PCC Population controls p10
- References p10

### Supplementary methods

#### Vaccination strategy in the Netherlands

The primary course vaccination campaign started in the beginning of 2021 in the Netherlands, prioritizing elderly, health care workers and at risk individuals. In adults, at the start of the Delta period (July 5th 2021) the coverage of the primary course was 47.0%, compared to 78.1% at the end (December 18th 2021). The vaccination coverage only marginally increased to 79.0% at the end of the Omicron inclusion period (May 31st 2022). The booster vaccination program prioritized the same groups at its onset and coverage increased from 47.4% to 65.6% during the Omicron period (January 3rd to May 31st 2022). The booster vaccine consisted of a single 0.5 mL dose mRNA-1273 90 'Moderna' or a single 0.3 mL dose BNT162b2 'Pfizer/BioNTech', which could be administered at least 3 months after a previous vaccination or SARS-CoV-2 infection.

#### Vaccination status definition

A complete primary course was defined as having received two doses mRNA-1273 90 'Moderna', BNT162b2 'Pfizer/BioNTech', or ChAdOx1 'Vaxzevria' vaccines, a heterogenous combination at least 14 days prior to a positive test or enrolment; or one dose of Ad26.COV2.S 'Jcovden' vaccine at least 28 days prior to a positive test or enrolment. Additionally, a primary course was defined as completed when a participant had a self-reported or confirmed SARS-CoV-2 infection more than 8 weeks prior to receiving one dose which itself was at least 14 days prior to a positive or enrolment. These definitions are in line with Dutch national guidelines for COVID-19 vaccination (1). Participants were partially vaccinated if they received a single dose of the two-dose vaccines or if they tested positive or enrolled within the immunization period. Participants were unvaccinated if they had received no dose at baseline. Finally, a participant was considered as 'boostered' when the primary course was completed and an additional dose was administered at least 7 days prior to a positive test or enrolment.

**Confounders and group comparisons**

Statistical analyses were performed according to our predefined study protocol and previous research (2, 3). Strata were formed with the predefined confounders age class (young (18-45 y.o.); middle (46-65 y.o.); old (>65 y.o.)), sex (female and male), level of education (low, median and high) and number of comorbidities (0, 1-2, >2) in order to perform indirect standardization of the primary outcomes. For all groups (cases and controls) and stratums a sample mean and variance was determined and subsequently pooled into an overall mean and variance of the mean. The overall mean was weighted to the relative size of the stratum (mean) and the square of the relative size (variance of the mean) within the Omicron cases. To compute 95% confidence intervals normal approximations from the pooled variances of the mean were used. Next, permutation tests on the sum statistics were performed to compare symptom prevalence and prevalence of severe symptoms between Omicron cases and controls, and Delta cases with the same aforementioned confounders. In the case of a stratum containing only participant from one group, the strata was omitted from testing, leading to lower sample sizes. Similarly, comparisons with permutations test were made between boostered Omicron cases and Omicron cases with a primary course and between Omicron case with a prior SARS-CoV-2 infection and Omicron cases without a suspected or confirmed SARS-CoV-2 infection.

**Scenarios for substitution of missing values at T3**

To assess the robustness of the complete case scenario, four alternative scenarios were implemented to assess robustness of the findings.

1. Carry forward; participants with missing values at T3 for reported symptoms and severity scores had these values carried forward from T0
2. Best case; participants with missing values at T3 for reported symptoms and severity scores had these values substituted as none present
3. Worst case; participants with missing values at T3 for reported symptoms and severity scores had these values substituted as all present
4. Multiple imputation; participants with missing values at T3 for reported symptoms and severity scores had these values imputed 20 times after which the data was pooled according to Ruben’s rules using age class, sex, level of education, number of comorbidities and participant group as predicting variables.

### R Packages

Multivariate Imputation by Chained Equations (MICE) package (4) for the multiple imputations and COIN package for the permutation tests were used (5).

### Supplementary Results

#### Table S1: Symptoms at baseline and three months in Delta and Omicron cases and test-negative and population controls.

| | Complete case analysis (n=21631) | | | | Complete case analysis (n=15391) | | | |
|---|---|---|---|---|---|---|---|---|
| | T0 | | | | T3 | | | |
| **Symptom** | **Delta cases, % (95 CI) (n=9486)** | **Omicron Cases, % (95 CI) (n=6136)** | **Test-negative controls, % (95 CI) (n=1931)** | **Population controls % (95 CI) (n=4078)** | **Delta Cases, % (95 CI) (n=6855)** | **Omicron Cases, % (95 CI) (n=4138)** | **Test-negative controls, % (95 CI) (n=1672)** | **Population controls % (95 CI) (n=2726)** |
| Fatigue | 58.4(57.4-59.3) | 57.5(56.3-58.7) | 40.9***(38.7-43.1) | 13.2***(12-14.3) | 29.3###(28.2-30.3) | 24.3(23.1-25.6) | 20.5**(18.4-22.5) | 15.6***(14.2-17) |
| Runny nose | 82.5(81.7-83.3) | 82.1(81.1-83) | 77.3***(75.3-79.3) | 16.5***(15.2-17.8) | 19.5###(18.5-20.4) | 16.2(15.1-17.3) | 24.6###(22.3-26.8) | 17.5(15.9-19) |
| Headache | 65.2##(64.2-66.2) | 63.1(61.9-64.2) | 45.8***(43.5-48.2) | 14.4***(13.3-15.6) | 18.7###(17.8-19.6) | 13.6(12.6-14.6) | 19.2###(17.2-21.2) | 15.5*(14-16.9) |
| Sneezing | 67.6(66.7-68.6) | 67.8(66.6-68.9) | 60.5***(58.2-62.8) | 12***(10.9-13.2) | 12.4(11.6-13.2) | 13.1(12-14.1) | 18.9###(16.8-20.9) | 13.2(11.7-14.7) |
| Coughing | 76.5***75.7-77.4) | 79.1(78.1-80.1) | 64.8***(62.6-67) | 9.7***(8.7-10.8) | 10.5(9.7-11.2) | 11.6(10.7-12.6) | 15.3##(13.5-17.2) | 12.8(11.3-14.3) |
| Dyspnoea | 29.5(28.6-30.4) | 28.3(27.2-29.4) | 20.8***(18.9-22.6) | 4.1***(3.4-4.8) | 15.9###(15-16.8) | 11.1(10.2-12.1) | 8.9*(7.5-10.4) | 6.7***(5.7-7.8) |
| Muscle pain/Joint pain (not sports related) | 48.6###(47.6-49.6) | 43.8(42.6-45) | 21.6***(19.6-23.5) | 6.0***(5.2-6.8) | 12.0###(11.2-12.8) | 9.5(8.7-10.4) | 9.9(8.4-11.5) | 9(7.8-10.3) |
| Difficulty concentrating | 15.2(14.4-15.9) | 14.7(13.9-15.6) | 8.4***(7.1-9.6) | 3.9***(3.3-4.5) | 13.8###(13-14.6) | 8.8(8-9.7) | 7.4(6-8.7) | 5.4***(4.5-6.3) |
| Sleep problems | 18.6(17.8-19.4) | 18.5(17.5-19.4) | 15**(13.4-16.5) | 7.9***(7-8.9) | 11.5###(10.7-12.3) | 8.8(7.9-9.6) | 9.4(7.9-10.9) | 6.6**(5.5-7.7) |
| Difficulties in busy environment | 12.1###(11.5-12.8) | 10.2(9.5-11) | 7.8***(6.6-9) | 4.6***(3.9-5.3) | 12.0###(11.3-12.8) | 8.4(7.5-9.2) | 6.0**(4.8-7.1) | 4.6***(3.8-5.5) |
| Sore throat | 50.2***(49.1-51.2) | 71.5(70.4-72.6) | 68.9**(66.8-71) | 7.3***(6.5-8.2) | 9.0(8.4-9.7) | 8(7.2-8.8) | 12.1###10.5-13.8) | 10.2###9-11.4) |
| Mucus from throat or nose | 32.2***(31.2-33.1) | 37.5(36.3-38.7) | 36(33.8-38.2) | 5.6***(4.8-6.5) | 6.7(6.1-7.4) | 6.3(5.6-7.1) | 7.3(5.9-8.6) | 6.7(5.6-7.8) |
| Ringing ears | 12.8(12.2-13.5) | 12.2(11.4-13.1) | 9.9**(8.5-11.3) | 5.6***(4.7-6.4) | 7.9##(7.2-8.5) | 6.2(5.5-7) | 7.6(6.2-8.9) | 6.2(5.2-7.3) |
| Problems with memory† | 2.6***2.3-3) | 4.4(3.9-4.9) | 2.1***(1.5-2.8) | 1.8***(1.4-2.2) | 8.9###(8.2-9.6) | 5.9(5.2-6.6) | 3.4***(2.5-4.3) | 3***(2.2-3.8) |
| Dizziness | 19##(18.2-19.8) | 16.9(16-17.9) | 8.9***(7.6-10.2) | 2.8***(2.2-3.4) | 5.8#(5.3-6.4) | 4.6(4-5.2) | 5.2(4-6.3) | 3.5(2.7-4.2) |
| Brainfog | 6.7*(6.2-7.2) | 7.6(7-8.3) | 3***(2.2-3.8) | 1.3***(0.9-1.7) | 5.9###(5.4-6.5) | 4.4(3.8-5) | 2.9*(2-3.8) | 1.9***(1.3-2.4) |
| Post exertional malaise | 7.5***(7-8.1) | 10.5(9.8-11.3) | 2.6***(1.9-3.3) | 0.8***(0.5-1.1) | 6.1###(5.5-6.7) | 4.1(3.5-4.7) | 3(2.1-3.9) | 2.1***(1.5-2.6) |
| General malaise | 42.2###(41.2-43.2) | 37.9(36.7-39.1) | 29.3***(27.1-31.4) | 2.6***(2.1-3.2) | 6.1###(5.5-6.6) | 3.9(3.3-4.5) | 6.3##5.1-7.6) | 4.5(3.6-5.4) |
| Chest pain or chest tightness | 15(14.3-15.7) | 14.2(13.4-15.1) | 5.8***(4.7-6.8) | 1.7***(1.2-2.2) | 5.3###(4.8-5.8) | 3.7(3.1-4.2) | 2.8(1.9-3.6) | 2.3*(1.7-2.9) |
| Altered stool | 14.9##(14.2-15.6) | 12.8(12-13.7) | 8.1***(6.9-9.4) | 2.2***(1.7-2.8) | 4.2(3.7-4.7) | 3.7(3.1-4.3) | 4.1(3.1-5.1) | 2.9(2.2-3.7) |
| Palpitations or tachycardia | 7(6.5-7.5) | 7.1(6.4-7.7) | 4.7***(3.7-5.6) | 2.1***(1.6-2.6) | 5.5###(4.9-6) | 3.7(3.2-4.3) | 3.9(2.9-4.9) | 2.8(2.2-3.5) |
| Abdominal pain | 9.2#(8.7-9.8) | 8.1(7.4-8.8) | 6.4*(5.2-7.5) | 2.9***(2.4-3.5) | 3.6(3.1-4) | 3.1(2.5-3.6) | 4.5#(3.4-5.6) | 2.3(1.8-2.9) |
| Nausea | 13.5(12.8-14.2) | 12.4(11.5-13.2) | 9.4***(8-10.8) | 2.4***(1.9-2.9) | 3.7#(3.2-4.1) | 2.9(2.4-3.4) | 4.0 (3-5) | 3(2.3-3.8) |
| Tingling or numbness | 4.5(4.1-5) | 4.3(3.8-4.8) | 2.1***(1.5-2.8) | 1.8***(1.3-2.2) | 4.0##(3.5-4.5) | 2.9(2.4-3.4) | 3.1(2.2-4) | 1.7*(1.2-2.3) |
| Sore eye | 6.1###(5.6-6.6) | 4.7(4.2-5.2) | 4.5(3.5-5.5) | 1.4***(0.8-1.9) | 2.3(1.9-2.6) | 2.4(1.9-2.9) | 2.1(1.4-2.8) | 1.1**(0.6-1.6) |
| Decreased appetite | 28.7###(27.8-29.6) | 20.1(19.1-21.1) | 11.2***(9.7-12.7) | 1.5***(1.1-1.9) | 2.7(2.4-3.1) | 2.2(1.8-2.7) | 2.7(1.8-3.5) | 1.8(1.3-2.3) |
| Loss of smell | 41.7###(40.7-42.7) | 11.8(11-12.6) | 5.2***(4.2-6.2) | 0.9***(0.5-1.3) | 12.1###(11.3-12.9) | 2.2(1.7-2.6) | 2.3(1.6-3.1) | 2.1(1.4-2.8) |
| Rash | 2.3(2-2.6) | 2.6(2.2-3) | 1.7(1.1-2.3) | 1.7**(1.2-2.2) | 2.4(2-2.8) | 2.2(1.7-2.6) | 2(1.2-2.7) | 2(1.4-2.6) |
| Depression | 1.1***0.9-1.3) | 1.9(1.5-2.2) | 2.3(1.7-2.9) | 1.7(1.3-2.1) | 2.4(2-2.8) | 2.2(1.7-2.6) | 3.1(2.3-4) | 1.5(1.1-2) |
| Shivers | 48.1###(47.1-49.2) | 43(41.8-44.2) | 23.2***(21.2-25.2) | 1.8***(1.4-2.2) | 4.8###(4.3-5.4) | 1.9(1.5-2.3) | 4.8###3.7-6) | 4.1###3.3-4.9) |
| Fever | 44.7###(43.6-45.7) | 38(36.7-39.2) | 20.3***(18.4-22.2) | 1.1***(0.8-1.5) | 2.3(2-2.7) | 1.8(1.4-2.2) | 2.3(1.5-3.1) | 3.3###2.6-4.1) |
| Menstruation problems | 1.2***(1-1.4) | 2.5(2.1-2.8) | 1.1***(0.6-1.5) | 1.5***(1.1-1.9) | 2.4(2-2.7) | 1.8(1.4-2.3) | 2(1.2-2.7) | 1.5(1-1.9) |
| Loss of taste | 36.5###(35.5-37.5) | 12(11.2-12.8) | 5.2***(4.2-6.3) | 0.6***(0.3-0.9) | 7.2###(6.5-7.8) | 1.7(1.3-2.1) | 1.5(0.9-2.1) | 1.4(0.8-1.9) |
| Anxiety | 1.9(1.6-2.2) | 2.1(1.7-2.4) | 1.6(1-2.1) | 1.6(1.2-2) | 2.1(1.7-2.4) | 1.7(1.3-2.1) | 2.4(1.7-3.2) | 1.6(1.1-2.1) |
| Neuralgia | 1.3***1.1-1.6) | 2.3(1.9-2.6) | 0.8***(0.4-1.3) | 0.9***(0.6-1.2) | 1.3(1-1.6) | 1.5(1.2-1.9) | 1.8(1.2-2.5) | 1.3(0.8-1.9) |
| Confusion | 2.7(2.3-3) | 2.5(2.1-2.9) | 1.4**(0.9-1.9) | 0.6***(0.3-0.9) | 2.5###(2.1-2.8) | 1.4(1-1.7) | 1.5(0.9-2.1) | 0.5**(0.2-0.7) |
| Nosebleed | 2.3***2-2.6) | 3.3(2.9-3.8) | 2.6(1.9-3.4) | 0.8***(0.4-1.1) | 1.5(1.2-1.8) | 1.2(0.9-1.5) | 1.8(1.1-2.5) | 1.1(0.7-1.6) |
| Earache | 10.9(10.2-11.5) | 10.7(10-11.5) | 7.6***(6.4-8.9) | 1.1***(0.7-1.4) | 1.8#(1.4-2.1) | 1.2(0.9-1.6) | 1.8(1.1-2.4) | 1.3(0.9-1.7) |
| Winter toes | 1.6(1.3-1.8) | 1.8(1.5-2.2) | 0.8**(0.4-1.2) | 0.9***(0.6-1.2) | 1.3###(1.1-1.6) | 0.6(0.4-0.8) | 1.2(0.6-1.7) | 1(0.6-1.4) |
| Vomiting | 2.4(2.1-2.7) | 2(1.7-2.4) | 1.7(1.1-2.3) | 0.6***(0.4-0.8) | 0.6(0.4-0.7) | 0.5(0.3-0.7) | 0.9(0.4-1.4) | 0.7(0.3-1) |
| New onset allergy† | 0**0-0.1) | 0.2(0.1-0.3) | 0(0-0) | 0.1(0-0.2) | 0.4(0.3-0.6) | 0.3(0.1-0.5) | 0.2(-0.1-0.4) | 0.0*(0-0) |
| **Severity cutoff scores** | | | | | | | | |
| CIS, subscale fatigue, ≥35 | 51.6###(50.7-52.6) | 45.7(44.5-46.8) | 32.3***(30.2-34.3) | 13.0***(11.9-14) | 25.6###(24.7-26.6) | 22.1(20.9-23.3) | 20.0 (18.1-22) | 14.1***(12.8-15.3) |
| CFQ, ≥44 | 9.7(9.1-10.2) | 9.8(9.1-10.5) | 11.2(9.8-12.7) | 7.3***(6.4-8.1) | 14.3###(13.5-15.1) | 11.8(10.9-12.8) | 10.6(9-12.1) | 6.6***(5.7-7.6) |
| mMRC, ≥1 | 16.8(16-17.5) | 15.6(14.7-16.5) | 11***(9.6-12.5) | 2.3***(1.8-2.8) | 12.2###(11.5-13) | 8.3(7.5-9.2) | 6.3*(5.1-7.6) | 4.6***(3.7-5.5) |
| SF-36 subscale bodily pain, ≤55 | 10.1##(9.5-10.7) | 8.4(7.7-9.1) | 9.1(7.8-10.3) | 5.4***(4.6-6.2) | 7.4(6.8-8.1) | 6.5(5.8-7.2) | 6.9(5.6-8.1) | 6.6(5.5-7.7) |

Standardised prevalence (95% confidence intervals) of participants with symptoms at T0 and T3 for cases, test-negative controls and population controls using complete analysis without substituting for missing values at T3. Negative values were truncated at 0.
* BH.adjusted p-value <0·05 compared to Omicron cases, prevalence higher in Omicron cases than in comparative group
** BH.adjusted p-value <0·01 compared to Omicron cases, prevalence higher in Omicron cases than in comparative group
*** BH.adjusted p-value <0·001 compared to Omicron cases, prevalence higher in Omicron cases than in comparative group
[#] BH.adjusted p-value <0·05 compared to Omicron cases, prevalence higher in comparative group than in Omicron cases
[##] BH.adjusted p-value <0·01 compared to Omicron cases, prevalence higher in comparative group than in Omicron cases
[###] BH.adjusted p-value <0·001 compared to Omicron cases, prevalence higher in comparative group than in Omicron cases

† Data collected on symptom at T0 only for part of the participants
CIS: Checklist Individual Strength
CFQ: Cognitive Failure Questionnaire
SF-36: SF-36 item Health Survey
mMRC: Modified Medical Research Council dyspnoea scale

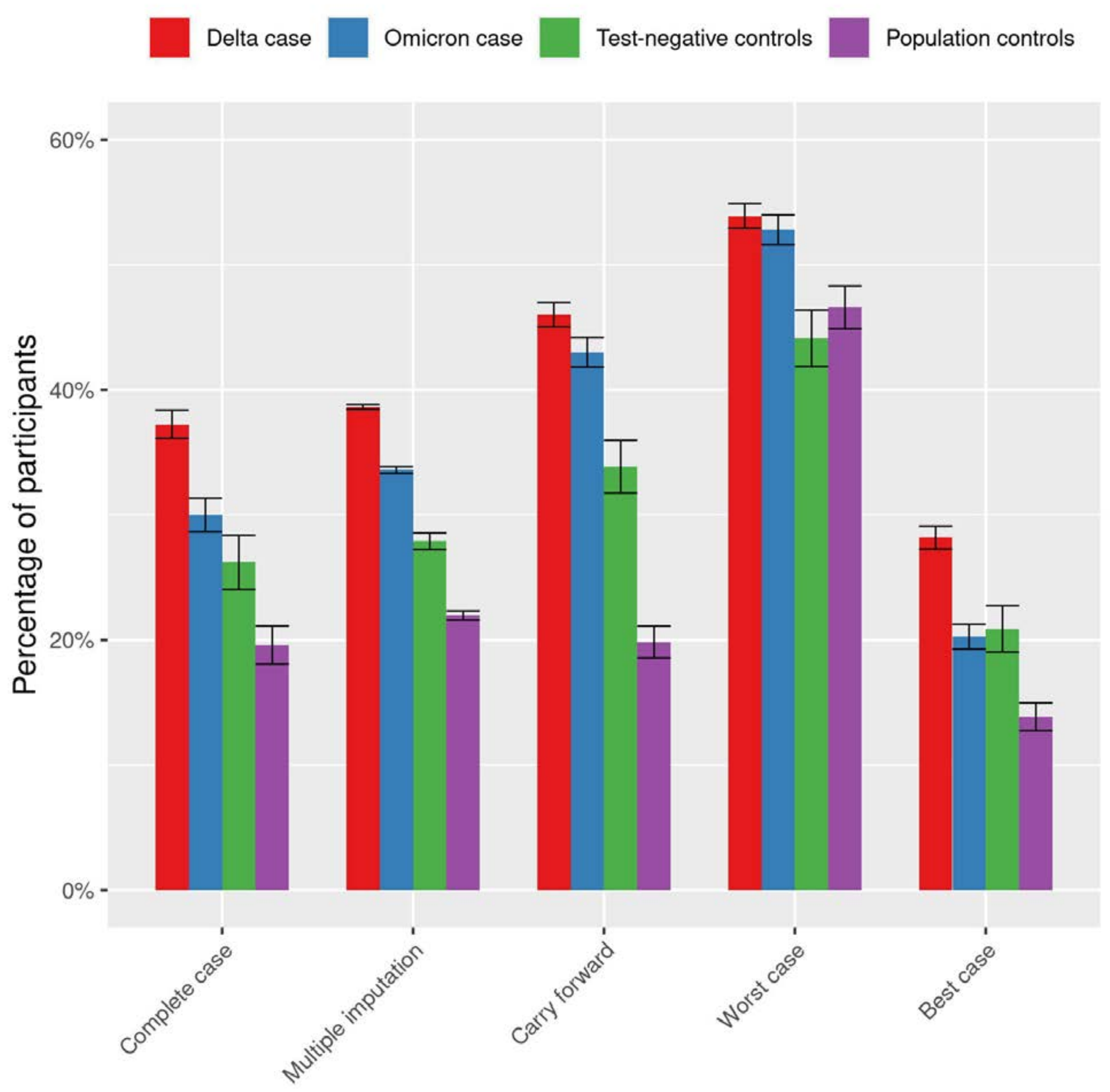


**Figure S1: prevalence of any symptom significantly elevated in cases compared to controls per scenario.** For complete case analysis scenario: n Delta cases: 6855, Omicron cases: 4138, Test-negative controls: 1672, Population controls: 2726; other scenarios: n Delta cases: 9486, Omicron cases: 6136, Test-negative controls: 1849 Population controls: 3945.

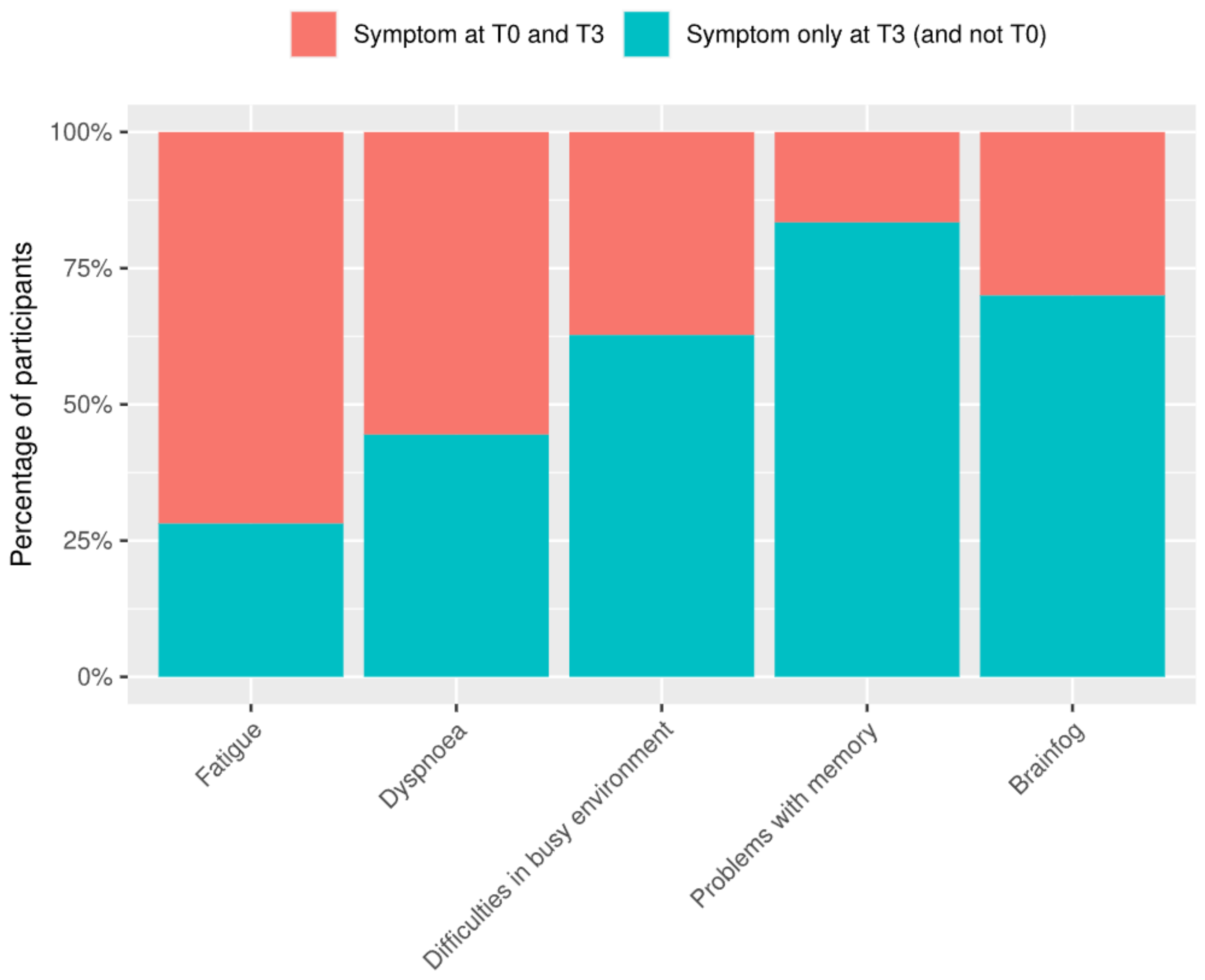


**Figure S2: Development of symptoms that are present at T3, i.e. present at T3 and T0 (persistent from T0 onwards), or present at T3 but not at T0 (late onset after T0) for cases per symptom**.

**Table S2. Demographics and acute illness at baseline for Omicron booster cases and primary course cases in the complete case scenario.**

| Complete case | Booster | Primary course |
|---|---|---|
| n | 2970 | 853 |
| Age, median [IQR] | 58.71 [46.97, 67.38] | 47.69 [34.65, 56.06] |
| Sex, % (n) | | |
| Female | 59.6 (1771) | 70.9 (605) |
| Male | 40.1 (1190) | 29.1 (248) |
| Other | 0.1 (4) | 0.0 (0) |
| Pregnancy, % (n) | 1.7 (9) | 3.0 (9) |
| BMI, mean (SD) | 25.98 (4.59) | 25.70 (4.94) |
| Smoking, % (n) | | |
| Current smoker | 3.6 (108) | 4.5 (38) |
| Former smoker | 28.0 (831) | 24.0 (205) |
| Never smoker | 66.5 (1974) | 68.9 (588) |
| Level of education, % (n) | | |
| Low | 3.1 (92) | 3.3 (28) |
| Medium | 31.6 (938) | 32.0 (273) |
| High | 65.3 (1940) | 64.7 (552) |
| Nr of comorbidities, % (n) | | |
| 0 | 42.4 (1258) | 50.4 (430) |
| 1-2 | 44.0 (1306) | 38.3 (327) |
| >2 | 13.7 (406) | 11.3 (96) |
| History of COVID-19, % (n) | 8.4 (250) | 17.6 (150) |
| Respiratory disease, % (n) | 18.0 (536) | 15.8 (135) |
| Hypertension, % (n) | 15.7 (466) | 9.6 (82) |
| Diabetes, % (n) | 4.3 (129) | 2.5 (21) |
| Cardiovasculair disease, % (n) | 2.9 (86) | 0.8 (7) |
| Medication use, % (n) | 74.0 (2199) | 75.8 (647) |
| Admitted to hospital, % (n)* | 0.1 (3) | 0.2 (2) |
| Number of symptoms at T0, median [IQR] | 8.00 [5.00, 11.00] | 9.00 [6.00, 13.00] |

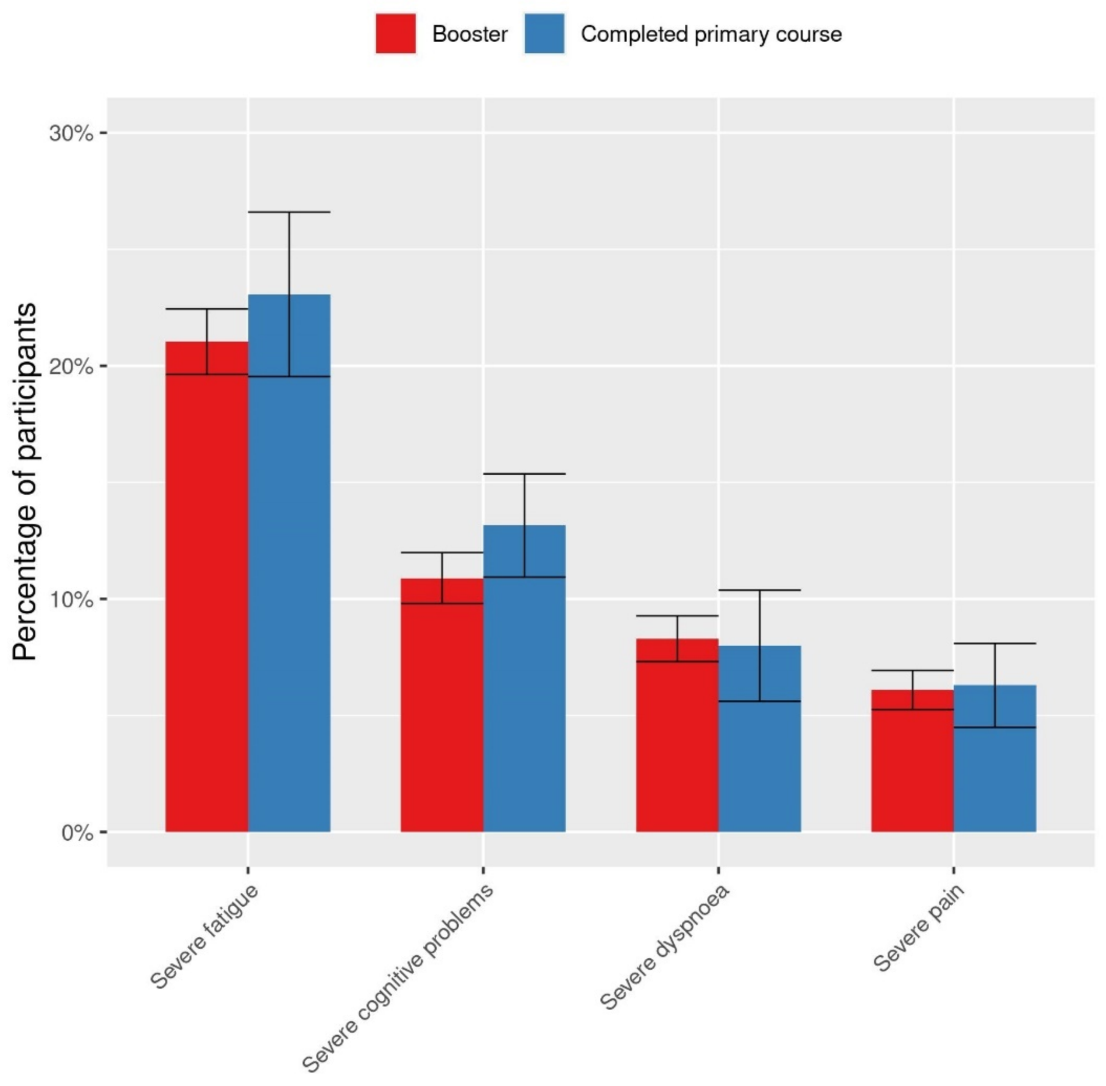


**Figure S3: Standardised prevalence (95% confidence intervals) of severity score cut-off values at in Omicron cases with a booster compared to Omicron cases with a completed primary course.** Complete case analysis without substituting for missing values at T3**.** Severe fatigue: Checklist Individual Strength (CIS), subscale fatigue ≥35, severe cognitive problems: Cognitive Failure Questionnaire (CFQ) ≥44, severe dyspnoea: modified Medical Research Council dyspnoea scale mMRC ≥1, severe pain: SF-36 subscale bodily pain ≤55.

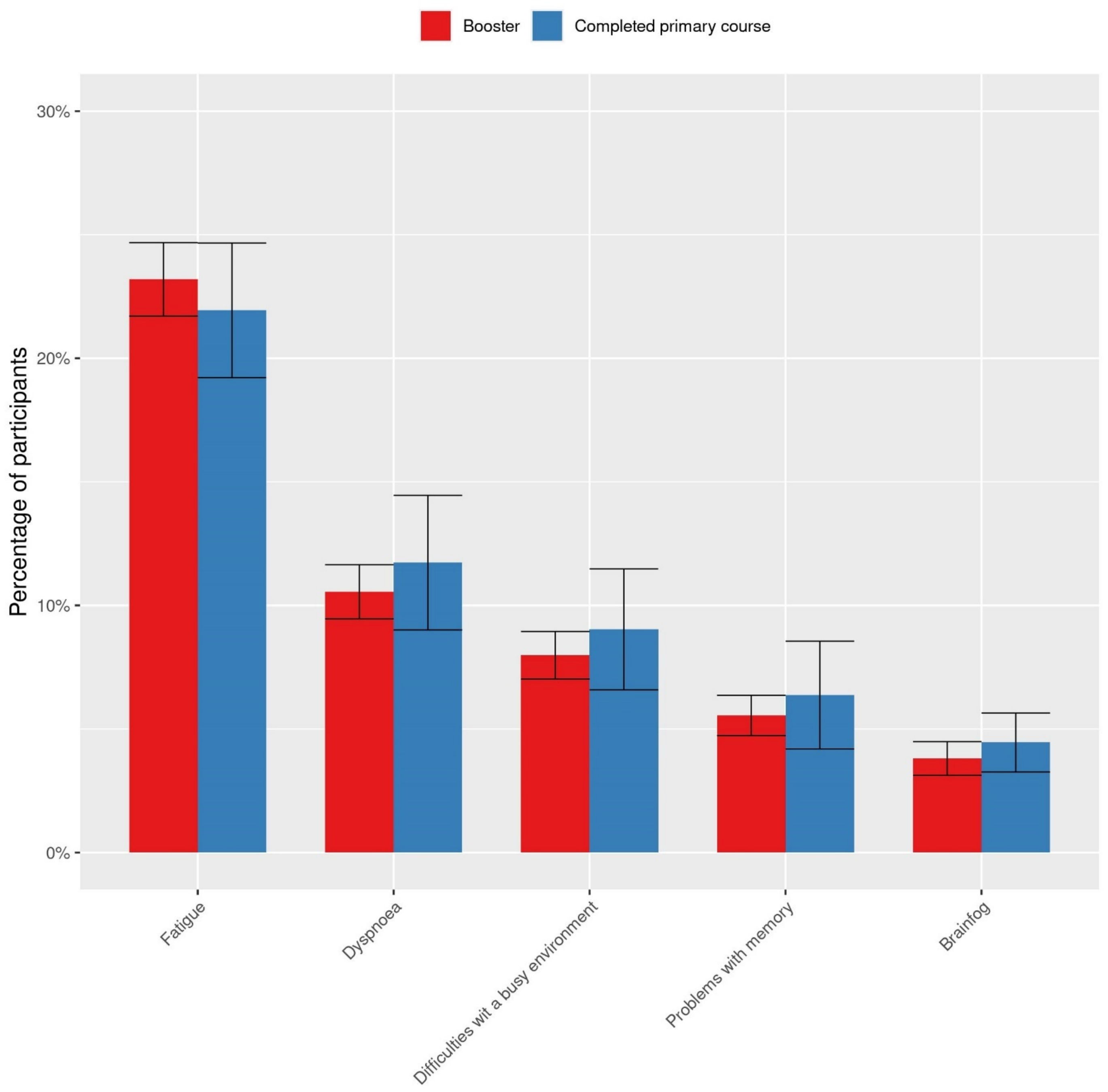


**Figure S4. Standardised prevalence (95% confidence intervals) of participants with symptoms at T3 for Omicron cases with a booster and with a primary course.** Complete analysis without substituting for missing values at T3**.**

**Table S3. Demographics and acute illness at baseline for Omicron cases with a reinfection and with a first infection in the complete case scenario**

| **Complete case** | **Omicron reinfection cases** | **Omicron first infection cases** |
|---|---|---|
| n | 453 | 3,685 |
| Age, median [IQR] | 52.01 [39.30, 60.47] | 56.33 [44.20, 66.21] |
| Sex, % (n) | | |
| Female | 66.9 (303) | 61.4 (2264) |
| Male | 32.9 (149) | 38.3 (1412) |
| Other | 0.0 (0) | 0.1 (4) |
| Pregnancy, % (n) | 0.8 (1) | 2.4 (18) |
| BMI, mean (SD) | 25.44 (4.61) | 25.97 (4.69) |
| Smoking, % (n) | | |
| Current smoker | 4.0 (18) | 3.9 (142) |
| Former smoker | 27.4 (124) | 26.9 (993) |
| Never smoker | 66.0 (299) | 67.1 (2472) |
| Level of education, % (n) | | |
| Low | 2.6 (12) | 3.3 (120) |
| Medium | 32.2 (146) | 32.0 (1178) |
| High | 65.1 (295) | 64.8 (2387) |
| Nr of comorbidities, % (n) | | |
| 0 | 42.2 (191) | 44.4 (1636) |
| 1-2 | 46.6 (211) | 42.2 (1555) |
| >2 | 11.3 (51) | 13.4 (494) |
| Respiratory disease, % (n) | 17,4 (720) | 16,6 (1137) |
| Hypertension, % (n) | 14,1 (584) | 12,0 (826) |
| Diabetes, % (n) | 3,7 (155) | 2,9 (202) |
| Cardiovasculair disease, % (n) | 18.8 (85) | 17.2 (635) |
| Medication use, % (n) | 74.0 (335) | 74.6 (2748) |
| Admitted to hospital, % (n)* | 2.0 (9) | 2.5 (93) |
| Vaccination status at T0, % (n) | | |
| Boostered | 60.7 (250) | 77.9 (2720) |
| Fully vaccinated | 36.4 (150) | 20.1 (703) |
| Partially vaccinated | 0.5 (2) | 0.5 (19) |
| Unvaccinated | 2.4 (10) | 1.4 (50) |
| Number of symptoms at T0, median [IQR] | 8.00 [5.00, 12.00] | 8.00 [5.00, 11.00] |

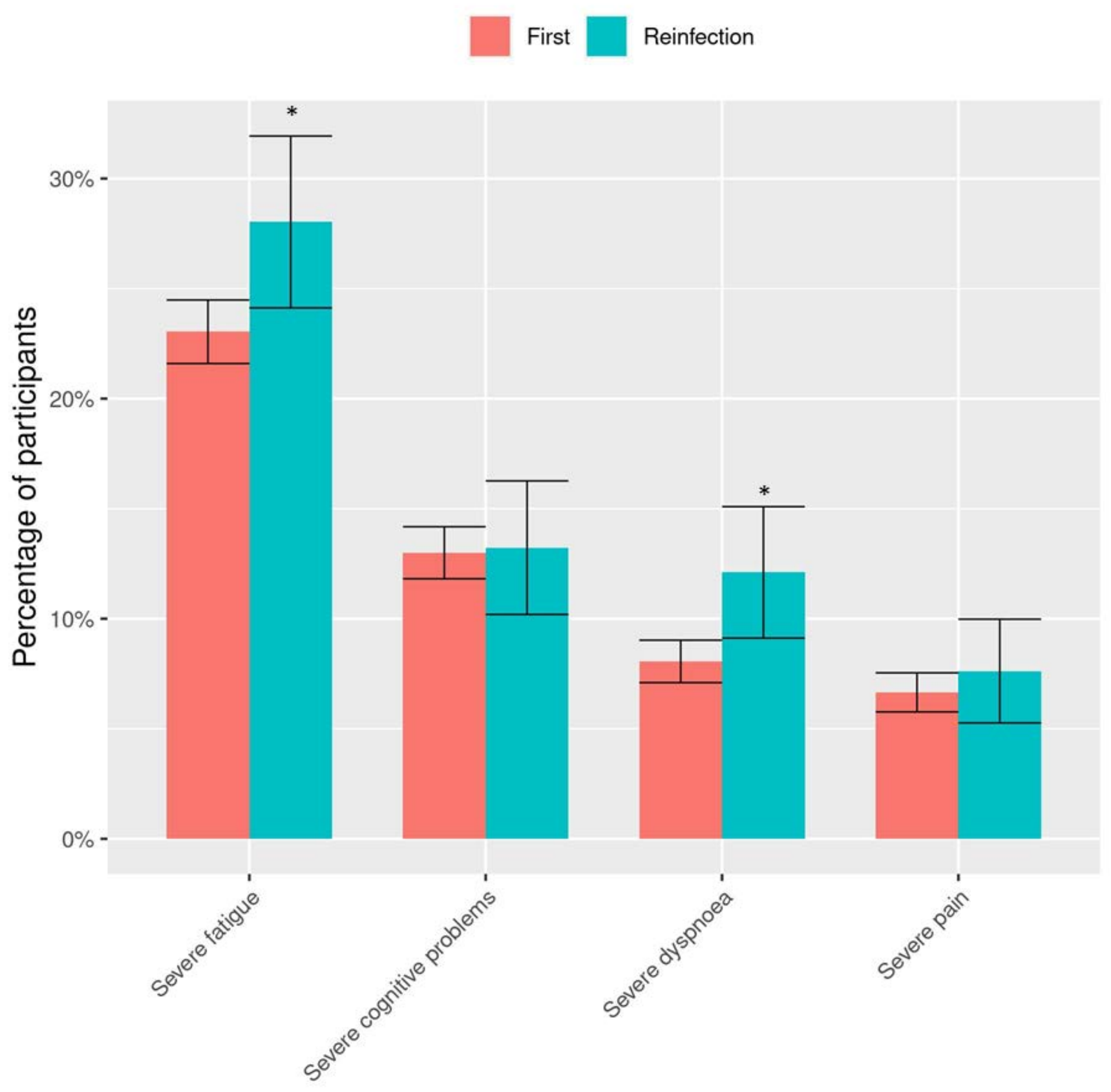


**Figure S5: Standardised prevalence (95% confidence intervals) of severity score cut-off values at in Omicron cases with first infection compared to Omicron cases with a reinfection.** Complete case analysis without substituting for missing values at T3. Severe fatigue: Checklist Individual Strength (CIS), subscale fatigue ≥35, severe cognitive problems: Cognitive Failure Questionnaire (CFQ) ≥44, severe dyspnoea: modified Medical Research Council dyspnoea scale mMRC ≥1, severe pain: SF-36 subscale bodily pain ≤55. *BH.adjusted p-value <0.05; **BH.adjusted p-value <0.01 ; ***BH.adjusted p-value <0.001 compared to Omicron cases with a first infection.

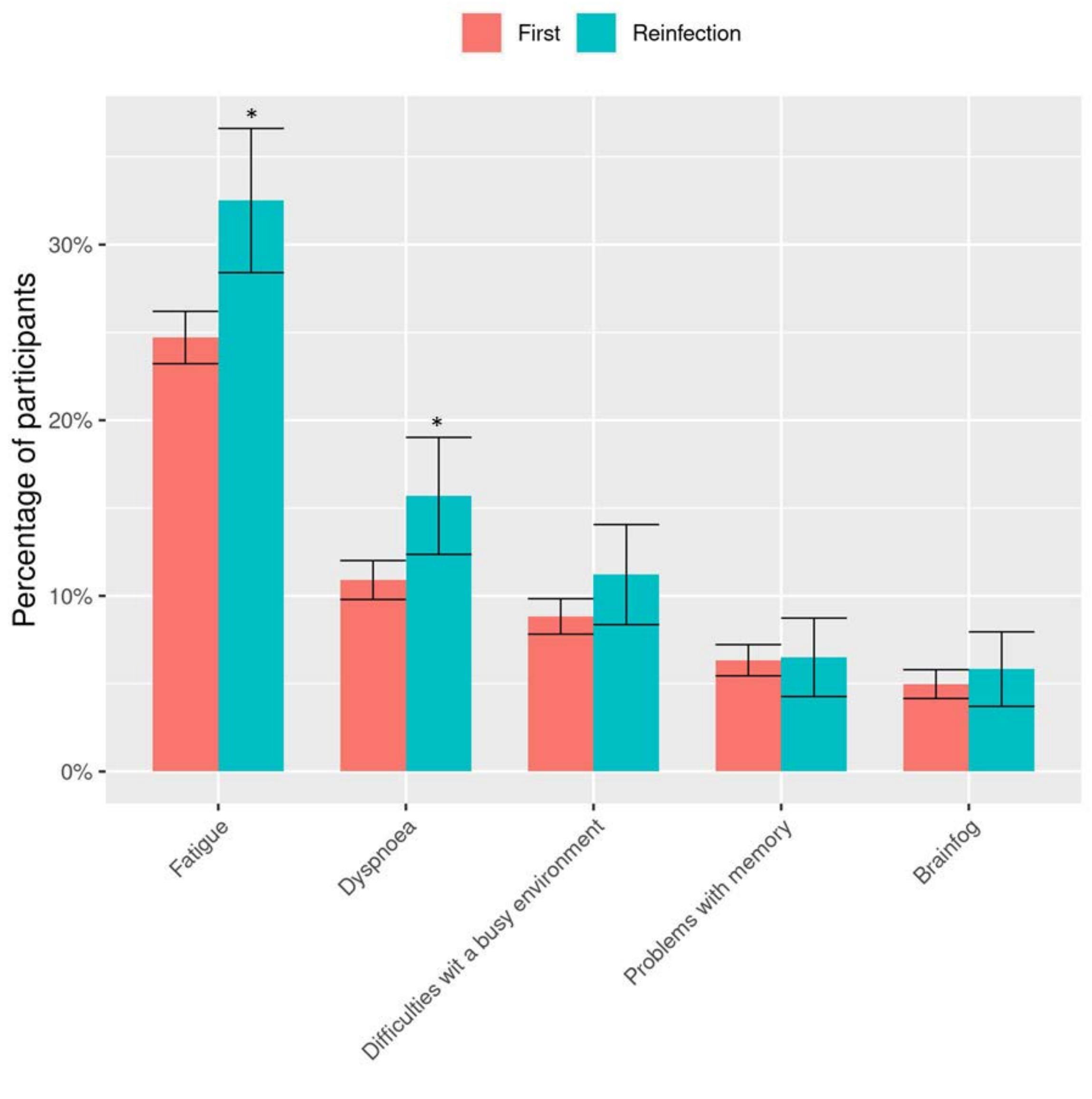


**Figure S6. Standardised prevalence (95% confidence intervals) of participants with symptoms at T3 for Omicron cases with a first infection and with a reinfection.** Complete analysis without substituting for missing values at T3**.** * BH.adjusted p-value <0·05 compared to first infection cases, prevalence higher in reinfected cases. *BH.adjusted p-value <0.05; **BH.adjusted p-value <0.01 ; ***BH.adjusted p-value <0.001 compared to Omicron cases with a first infection.

**Table S4. Severity score for CIS fatigue, CFQ score, SF-36 bodily pain compared between non-PCC Omicron cases and non-PCC Population controls**

| Severity score | non-PCC Omicron cases | non-PCC Population controls | p.BH-value |
|---|---|---|---|
| CIS fatigue score | 18.7 (18.4 - 19.1) | 17.9 (17.5 - 18.3) | 0,0024 |
| CFQ score | 23.6 (23.2 - 24.0) | 22.8 (22.3 - 23.4) | 0,00459 |
| SF-36 bodily pain score | 89.7 (89.2 - 90.3) | 88.8 (88.0 - 89.6) | 0,0384 |

### References

1. Rijksinstituut voor Volksgezondheid en Milieu (RIVM). Guidelines COVID-19 vaccination. 2023 [Available from: https://lci.rivm.nl/richtlijnen/covid-19-vaccinatie.
2. Mutubuki EN, van der Maaden T, Leung KY, Wong A, Tulen AD, de Bruijn S, et al. Prevalence and determinants of persistent symptoms after infection with SARS-CoV-2: protocol for an observational cohort study (LongCOVID-study). BMJ Open. 2022;12(7):e062439.
3. van der Maaden T, Mutubuki EN, de Bruijn S, Leung KY, Knoop HA-O, Slootweg J, et al. Prevalence and severity of symptoms 3 months after infection with SARS-CoV-2 compared to test-negative and population controls in the Netherlands. LID - jiac474 [pii] LID - 10.1093/infdis/jiac474 [doi]. (1537-6613 (Electronic)).
4. van Buuren SG-O, K. mice: Multivariate Imputation by Chained Equations in R. Journal of Statistical Software. 2011;45(3):1-67.
5. Hothorn TH, K.; van de Wiel, M.A.V.; Zeileis, Al. Implementing a Class of Permutation Tests: The coin Package. J Stat Softw. 2008;28(8):1-23.
